# Protein identification for stroke progression via Mendelian Randomization in Million Veteran Program and UK Biobank

**DOI:** 10.1101/2024.01.31.24302111

**Authors:** Andrew Elmore, Nimish Adhikari, April E Hartley, Hugo Javier Aparicio, Dan C. Posner, Gibran Hemani, Kate Tilling, Tom R Gaunt, Peter Wilson, JP Casas, John Michael Gaziano, George Davey Smith, Lavinia Paternoster, Kelly Cho, Gina M Peloso

**Affiliations:** NIHR Bristol Biomedical Research Centre, University Hospitals Bristol and Weston NHS Foundation Trust and University of Bristol; MRC Integrative Epidemiology Unit (IEU), Bristol Medical School, University of Bristol; Veteran’s Affairs Healthcare System, Boston, MA; Department of Biostatistics, Boston University School of Public Health, Boston, MA; Department of Neurology, Boston University Chobanian & Avedisian School of Medicine, Boston, MA; Boston Medical Center, Boston, MA; Division of Aging, Brigham and Women’s Hospital, Harvard Medical School; Atlanta VA Medical Center, Atlanta, GA

**Keywords:** Progression, Prognosis, Stroke, AIS, MACE, Mendelian Randomization, GWAS

## Abstract

**Background:** Individuals who have experienced a stroke, or transient ischemic attack, face a heightened risk of future cardiovascular events. Identification of genetic and molecular risk factors for subsequent cardiovascular outcomes may identify effective therapeutic targets to improve prognosis after an incident stroke.

**Methods:** We performed genome-wide association studies (GWAS) for subsequent major adverse cardiovascular events (MACE) (N_cases_=51,929, N_cntrl_=39,980) and subsequent arterial ischemic stroke (AIS) N_cases_=45,120, N_cntrl_=46,789) after first incident stroke within the Million Veteran Program and UK Biobank. We then used genetic variants associated with proteins (pQTLs) to determine the effect of 1,463 plasma protein abundances on subsequent MACE using Mendelian randomization (MR).

**Results:** Two variants were significantly associated with subsequent cardiovascular events: rs76472767 (OR=0.75, 95% CI = 0.64-0.85, p= 3.69x10^-08^) with subsequent AIS and rs13294166 (OR=1.52, 95% CI = 1.37-1.67, p=3.77x10^-08^) with subsequent MACE. Using MR, we identified 2 proteins with an effect on subsequent MACE after a stroke: *CCL27* (effect OR= 0.77, 95% CI = 0.66-0.88, adj. p=0.05), and *TNFRSF14* (effect OR=1.42, 95% CI = 1.24-1.60, adj. p=0.006). These proteins are not associated with incident AIS and are implicated to have a role in inflammation.

**Conclusions:** We found evidence that two proteins with little effect on incident stroke appear to influence subsequent MACE after incident AIS. These associations suggest that inflammation is a contributing factor to subsequent MACE outcomes after incident AIS and highlights potential novel targets.

## Introduction

Stroke remains a significant public health concern worldwide. With its potential to cause profound disabilities and mortality, it necessitates continued research efforts to unravel its multifaceted aetiology, identify modifiable risk factors, and develop effective therapeutic interventions.

Arterial ischemic stroke (AIS) accounts for approximately 85% of all stroke cases and arises from occlusion of cerebral blood vessels, leading to inadequate perfusion and a subsequent ischemic cascade(1). Through the study of incident stroke events, modifiable factors such as hypertension, diabetes, dyslipidaemia, atrial fibrillation, obesity, and lifestyle behaviours have been identified, which may offer promising targets for prevention (2). Whether targeting the same factors offer avenues for effective treatment after the incident event is unclear.

Genome wide association studies (GWAS) are usually performed on disease status for incident events but expanding them to subsequent events could provide us with novel biological insights about stroke progression, which may be more relevant for drug identification opportunities(3). GWAS of stroke incidence have previously observed 32 loci associated with stroke and stroke subtypes(5) with a recent study adding 5 more novel loci for stroke incidence(5). GWAS of disease progression can provide genetic risk factors that may be independent of the incident event. Since GWAS of disease progression include only individuals with incident disease, this can lead to the statistical problem of collider bias (or index-event bias), where shared confounders between incident and subsequent events can uncover spurious associations and biased estimates of effects, even amongst genetic risk factors(3)

Mendelian randomization (MR) is an established statistical method that uses genetic variants to assess putative causal relationships between genetically proxied protein abundance on incident AIS and subsequent AIS and MACE(6). The main advantage MR has over traditional observational epidemiological methods is that MR can imply causality between an exposure and an outcome because it is less liable to common epidemiological biases, such as confounding and reverse causality. For biases that MR does not account for, sensitivity analyses can assess whether results are robust. One such method is colocalization, which is used to identify if a genetic variant is shared by two traits and is a necessary condition for causality(7).

In this study, we perform GWAS of subsequent AIS and MACE after incident AIS in the Million Veteran Program and UK Biobank stratified by ancestry and meta-analysed across ancestries. We then use our subsequent events GWAS to perform MR for plasma protein abundances using pQTLs from UK Biobank Pharma Proteomics Project. Our genetic study aims to mimic a stroke prevention trial where recruitment into the trial is based on having a primary stroke event.

## Methods

### Genome Wide Association Studies

#### Phenotype Definitions

Incident stroke was defined as any diagnosis of AIS or transient ischaemic attack (TIA) using hospital linked data. People who experienced their initial stroke more than one year prior to recruitment were excluded from our stroke phenotype. Specific International Classification of Disease (ICD) codes used for both MVP and UKB can be found in the supplementary information.

Subsequent AIS/TIA was defined as any secondary diagnosis of AIS/TIA at least 90 days after the incident diagnosis, to avoid recoding of the primary event, and would be considered events after the acute phase of an incident AIS/TIA. Individuals who did not survive at least thirty days after their incident stroke diagnosis were excluded from analyses of subsequent outcomes, to emulate a target clinical trial. Subsequent MACE was defined as any subsequent stroke, myocardial infarction (MI), or death due to atherosclerotic cardiovascular disease (ASCVD), with the first event that happens after 90 days used to construct the MACE phenotype. Vascular disease events occurring before or after the initial stroke were excluded, but events greater than 90 days post stroke were included.

#### UK Biobank (UKB)

UKB is a prospective cohort study with over 500,000 participants aged 40-69 (average 56.5) years when recruited in 2006-2010 and 54% of participants are women(8). Information on the genotype imputation, quality control and GWAS is available in the Supplementary Information.

#### Million Veteran Program (MVP)

MVP is a continually growing cohort of over 850,000 participants by 2021(9), 8% women, with an average age of 61.9 years(10). Information on the genotyping, imputation, quality control and GWAS is available in the Supplementary Information.

#### Collider Bias Sensitivity Analysis and Correction

To perform a correction for collider bias for subsequent stroke we used Slope-Hunter(11), a method that uses a mix of thresholding and mixed model clustering to quantify the bias and present a corrected estimate of the progression effect of a subsequent stroke. Slope-Hunter assumes that SNPs can be divided into clusters based on their causal relationship with incident and subsequent events and uses SNPs associated with the incident event only to provide an estimate of the bias correction factor for the study, hence is more robust to the correlation between incident and subsequent events. However, when investigating specific SNPs and their associated regions, collider bias correction may only be necessary if there is an association of the variant with incident AIS to begin with. For that reason, we have compared Slope-Hunter adjusted results with non-Slope-Hunter adjusted results, as well as compared the results with the associated region in the incident GWAS. Each Slope-Hunter calculation was performed for each specific ancestry as the collider bias may behave differently in each subset of data.

We used the Slope-Hunter method with a default p-value threshold of 0.001 to correct the summary statistics for further analyses. We used the 1000genomes reference panel for clumping matched by ancestry group, with an r^2^ threshold of 0.1.

#### Expected vs. Observed Replication

To determine whether the GWAS results of subsequent stroke are different from incident stroke, we used the approach described in Okbay et. al(12), that determines replication performance by accounting for differential power. Here we used the approach to determine the extent to which incidence stroke GWAS hits are replicated in subsequent stroke, compared against the power-adjusted expected replication rate.

### Multi-Ancestry Comparison and Meta-analysis

Two meta-analyses were performed. First, European only meta-analysis was performed across UKB and MVP. Secondly a meta-analysis of all individuals, including each ancestry from MVP (European, African and Hispanic) and Europeans from UKB was conducted using a fixed-effects model. We completed this meta-analysis for both original and Slope-hunter adjusted results, and compared the results. Both meta-analyses and heterogeneity score calculations were performed using the software METAL(13). The Cochran’s Q-Statistic was used to test for heterogeneity between ancestries(13). We set our genome-wide significance threshold to 5x10^-08^.

We ran tissue expression analysis on subsequent stroke states using Functional Mapping and Annotation of GWAS (FUMA)(14), including MAGMA(15) Tissue Expression Analysis to investigate if there were any significant correlations between the subsequent GWAS and tissue expression.

### Mendelian Randomization against Protein Abundance

Using the meta-analysed GWAS results and existing available protein quantitative trait loci (pQTL) data sets, we performed MR for each outcome with a panel of 1,463 plasma proteins as potential causal risk factors. Measuring proteins at population scale could help discover novel clinical biomarkers and improve fine-mapping of causal genes linked to complex diseases(16). To account for multiple testing, p-values were adjusted using false discovery rate (FDR), and are subsequently reported as adjusted p.

pQTLs were extracted from pQTL studies from UK Biobank Pharma Proteomics Project (UKB-PPP) (54,306 participants, 1,463 proteins, Olink platform) (16). To ensure the robustness of the instruments (pQTLs), we attempted to replicate the MR results using 3 independent pQTL datasets; Atherosclerosis Risk in the Community (ARIC, European and African ancestry) (9,084 participants, 4,657 proteins, SOMAScan platform)(17), deCODE (35,559 participants, 4,907 proteins, SOMAScan platform) (18), and INTERVAL (3,301 participants, 3,622 proteins, SOMAScan platform) (19) MR for subsequent stroke MACE and AIS were performed on the multi-ancestry meta-analysis. MR for incident stroke AIS was also performed on the largest known stroke GWAS published(4). MR analyses were performed using the ‘TwoSampleMR’ R package(20).The pQTL data sets were meta-analysed across ancestries (16,18,19). We used a two-sample MR framework to estimate the putative causal effect of genetically proxied protein abundance to incident and subsequent stroke. MR estimates were generated using the Wald ratio method for instruments consisting of single SNPs, which included all of our instruments.

MR relies on three assumptions for identifying a putative causal effect(21), the genetic instrument should: 1) associate with the exposure (relevance), 2) have no shared causal factors with the outcome (independence), and 3) solely influence the outcome through the impact of the risk factor of primary concern (exclusion restriction). The relevance assumption was tested by generating the F-statistic for each instrument, where an F-statistic > 10 is evidence against weak instrument bias(22). The exclusion restriction assumption is difficult to assess with single SNP instruments, as is common for molecular traits. Therefore, we additionally performed colocalization. Finally, to explore if there was any evidence of heterogeneity of effects between genetic ancestries, for SNPs used in MR, we compared the associations with the outcomes across ancestries.

#### Colocalization

Colocalization is a phenomenon whereby genetic factors at a particular locus are shared between two or more traits. The package *coloc* was used to assess whether two association signals are consistent with a shared causal variant(7). We assessed the posterior probabilities of if the analysed SNPs share the same causal variant (known as H4)(7), where H4 ≥ 80% indicates strong evidence, and 80% > H4 ≥ 60% indicates moderate evidence of colocalization.

#### Collider Bias Analysis and Correction

We tested whether SNPs used in the MR were associated with the published stroke incidence to determine the potential for collider bias and ran MR against both the uncorrected meta-analysed GWAS as well as the Slope-Hunter adjusted meta-analysed GWAS, as explained previously.

### Compare Results Against Known Druggable Targets

Once significant SNPs from GWAS results and proteins from MR results were identified, we cross-referenced these with known existing SNPs, as well as existing literature around stroke onset and progression. Finally, we compared the results from the pQTL MR against known druggable targets from Open Targets(23).

## Results

### Genome Wide Association Studies

After exclusions based on ancestry and relatedness, 93,422 individuals who had an incident stroke across the UKB and MVP were analysed (86,237 for MVP, 7,185 for UKB), among which 51,929 had subsequent MACE and 45,120 has subsequent AIS. Stroke cases were older, more commonly male, with a higher proportion of smokers and individuals with hypertension, type 2 diabetes, anti-hypertensive use, and lipid-lowering medication use than individuals who had never experienced an AIS (**Table S1**).

There were no genome-wide significant associations in the multi-ancestry meta-analysis for subsequent AIS or MACE events (**Figure S1 and S2**), but we did observe 2 genome-wide significant (p<5x10^-08^) genetic variants in specific ancestry analyses: rs76472767 near gene *RNF220* on chromosome 1 in the AFR GWAS for subsequent MACE (slope-hunter corrected p = 3.69x10^-08^) and rs13294166 near gene *LINC01492* on chromosome 9 in the AFR GWAS for subsequent AIS (uncorrected p=3.77x10^-08^) (**Figure 2, Table S2**). For these two associations, we compared the results before and after Slope-Hunter correction as well as with the results in the incident AIS. We observed that none of the significantly associated variants were associated with incident AIS, and therefore the Slope-Hunter correction for collider bias may not have been necessary and the uncorrected results may be considered unbiased (**Figure 2**). However, the Slope-Hunter correction for collider bias may lead to slight differences in the results.

**Figure 1:**
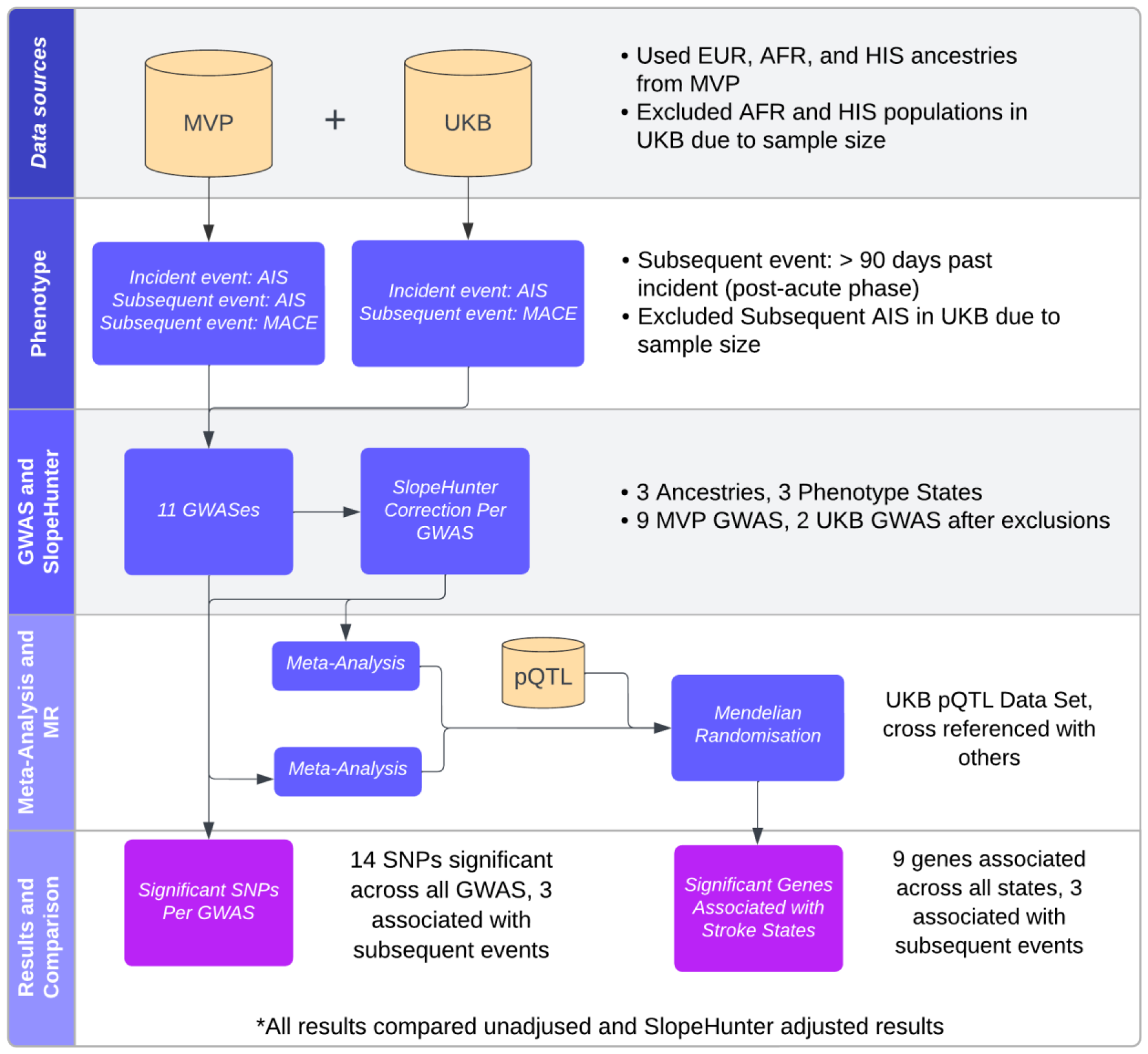
Flowchart of the methodological processes for analysing stroke data. MVP: Million Veterans Program; UKB: United Kingdom Biobank; AIS Acute Ischemic Stroke; MACE: Major Acute Cardiovascular Events; GWAS: Genome Wide Association Study; SNP: Single Nucleotide Polymorphism; pQTL: Protein Quantitative Trait Loci; MR: Mendelian Randomization

**Figure 2:**
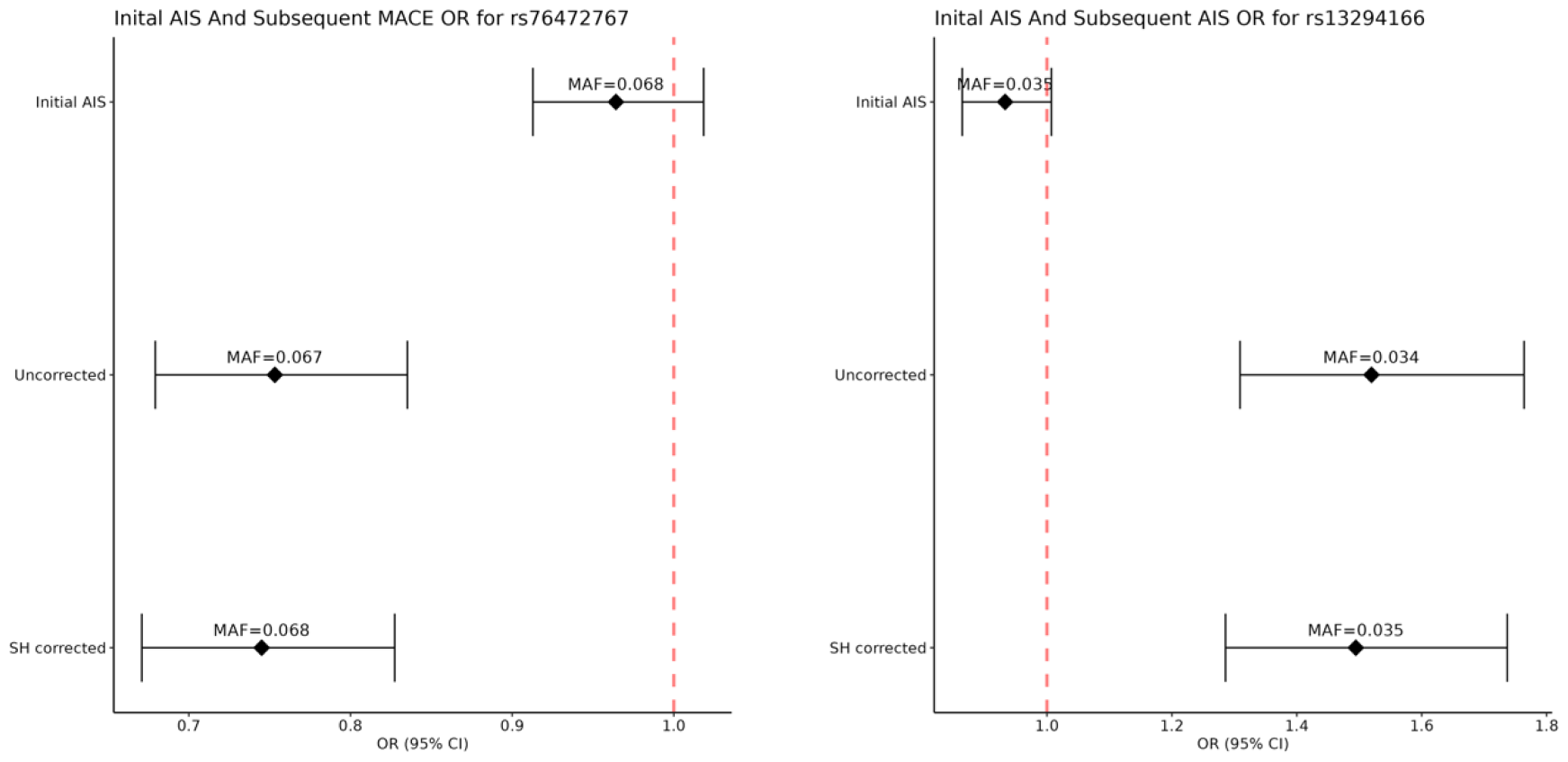
Forest Plots of Initial AIS and corrected and uncorrected subsequent events for the two genome wide significant SNPs. Generalized linear mixed effect regression models were used to test for associations between minor alleles and Subsequent MACE in stroke patients, adjusted for the first 10 genetic principal components. AIS: Acute Ischemic Stroke; MACE: Major Acute Cardiovascular Events; SH corrected: Result corrected by Slope-Hunter; OR: Odds ratio; SNP: Single Nucleotide Polymorphism

We tested for tissue enrichment of subsequent stroke GWAS signals using expression data with MAGMA in FUMA. However, no tissues were expressed above any statistically significant threshold (**Figure S4**).

#### Expected vs. Observed Replication

We sought to determine if the genetic factors for incident stroke were also relevant for subsequent stroke. Of the 91 SNPs previously reported to associate with incident stroke, we observed that 77 replicated in our incidence GWAS at p < 0.05 (91 expected given the power difference). By contrast, our subsequent MACE GWAS replicated only 33 (compared to 82 expected, p_diff_ = 3x10^-35^), suggesting there is overlapping, but also distinct genetic aetiology of incident stroke and subsequent MACE. This pattern was consistent when using collider bias corrected results (**Table S3**).

### Mendelian Randomization against pQTL Data

The subsequent AIS MR results were similar to subsequent MACE, but due to lower sample size, had larger p-values and wider confidence intervals, therefore, we focus our MR study on the subsequent MACE results (full results in **Table S4**). We observed 6 genes for incident stroke and 2 genes for subsequent stroke that have a significant MR result (adj. p<0.05) and supporting colocalization evidence (PP H4>60%) (**Table 1**). For all 6 genes, all MR results are based on single instrumental variant since only one cis pQTL was available and the Wald ratio was used.

**Table 1:** MR and colocalization results from UKB-PPP pQTL dataset against both incident and subsequent MACE. MR: Mendelian randomization; MACE: Major Acute Cardiovascular Events; pQTL: Protein Quantitative Trait Loci; MR: Mendelian randomization; UKB-PPP: United Kingdom Biobank Pharma Proteomics Project; coloc h4: Posterior probability that the analysed SNPs in the region share one common causal variant.

| Protein | Incident MR OR | Incident MR SE | Incident MR Adjusted P | Incident coloc h4 | Subsequent MR OR | Subsequent MR SE | Subsequent MR Adjusted P | Subsequent coloc h4 |
| --- | --- | --- | --- | --- | --- | --- | --- | --- |
| **Proteins associated with incident stroke** |  |  |  |  |  |  |  |  |
| CST6 | 1.079 | 0.0183 | 0.00489 | 85.8% | 0.958 | 0.0431 | 0.967 | N/A |
| FGF5 | 1.063 | 0.00837 | 1.55e-10 | 94.9% | 1.022 | 0.0171 | 0.962 | N/A |
| FURIN | 1.134 | 0.0337 | 0.0173 | 95.1% | 1.016 | 0.0592 | 0.987 | N/A |
| GRK5 | 0.849 | 0.0258 | 5.19e-08 | 99.9% | 0.959 | 0.0461 | 0.967 | N/A |
| MMP12 | 0.944 | 0.0117 | 1.30e-04 | 88.8% | 0.973 | 0.0216 | 0.962 | N/A |
| SCARA5 | 0.936 | 0.0182 | 0.0268 | 76.6% | 0.990 | 0.0358 | 0.987 | N/A |
| **Proteins associated with subsequent MACE** |  |  |  |  |  |  |  |  |
| CCL27 | 0.961 | 0.0395 | 0.831 | 0.7% | 0.770 | 0.0565 | 0.00618 | 68.3% |
| TNFRSF14 | 1.001 | 0.00126 | 0.949 | 0.4% | 1.419 | 0.0923 | 0.0469 | 81.6% |

#### Incident pQTL Results

We identified 6 proteins (CST6, FGF5, FURIN, GRK5, MMP12, SCARA5) with evidence for a putative causal effect on incident AIS (adj. p<0.05). However, none of these showed evidence for a putative causal effect on subsequent MACE (**Table S4**). All except SCARA5 showed very strong evidence for colocalization, while SCARA5 showed moderate evidence of colocalization (**Figure S5**).

#### Subsequent MACE pQTL Results

Two proteins (CCL27 and TNFRSF14) showed evidence for a putative causal effect on subsequent stroke (adj. p<0.05, **Table 1**). Neither of these proteins showed a putative causal effect on incident stroke. Genetically predicted higher levels of CCL27 showed evidence of a protective effect against subsequent stroke (OR=0.77, 95% CI = 0.66, 0.88). In contrast, higher predicted TNFRSF14 levels increased risk of subsequent stroke (OR=1.419, 95% CI =1.24, 1.60). After collider bias correction using Slope-Hunter, the MR results for CCL27 and TNFRSF14 were not significantly affected; CCL27: 0.77 (95% CI = 0.65, 0.89), TNFRSF14: 1.419 (95% CI = 1.24, 1.60). (**Table S4**). However, as these specific variants were not associated with incident disease (**Table 1**) the potential for collider bias was minimal. Both proteins implicated have a role in inflammation(24,25).

There was evidence for colocalization of CCL27 and TNFRSF14 protein and subsequent MACE (**Table 1, Table S5, Figure S6 and S7**). The assignment of the remaining probability mostly to H1 (representing association only with protein trait and not stroke outcome) suggests this analysis has limited power, rather than suggesting there are independent effects that don’t colocalise (H3).

#### Verification of pQTL instruments using other datasets

To verify that the protein instruments identified in UKB-PPP were valid, we replicated the MR results using 3 other independent pQTL data sets (ARIC, deCODE and INTERVAL). For 5 of the 9 significant MR results, MR using independent pQTL data sets showed consistent putative causal effects (**Table S6**). Due to the differing power of the pQTL data sets, only pQTLs were filtered for having F-statistic value above 10 (**Table S7**).

#### Multi-Ancestry Comparison of MR Results and Meta-analysis

There is little evidence that the 3 proteins reported as having putative causal effects on subsequent stroke have different putative causal effects across the three ancestries tested. However, this is primarily due to the very wide confidence intervals (and small sample sizes) within Hispanic and African subgroups (**Figure 3**).

**Figure 3:**
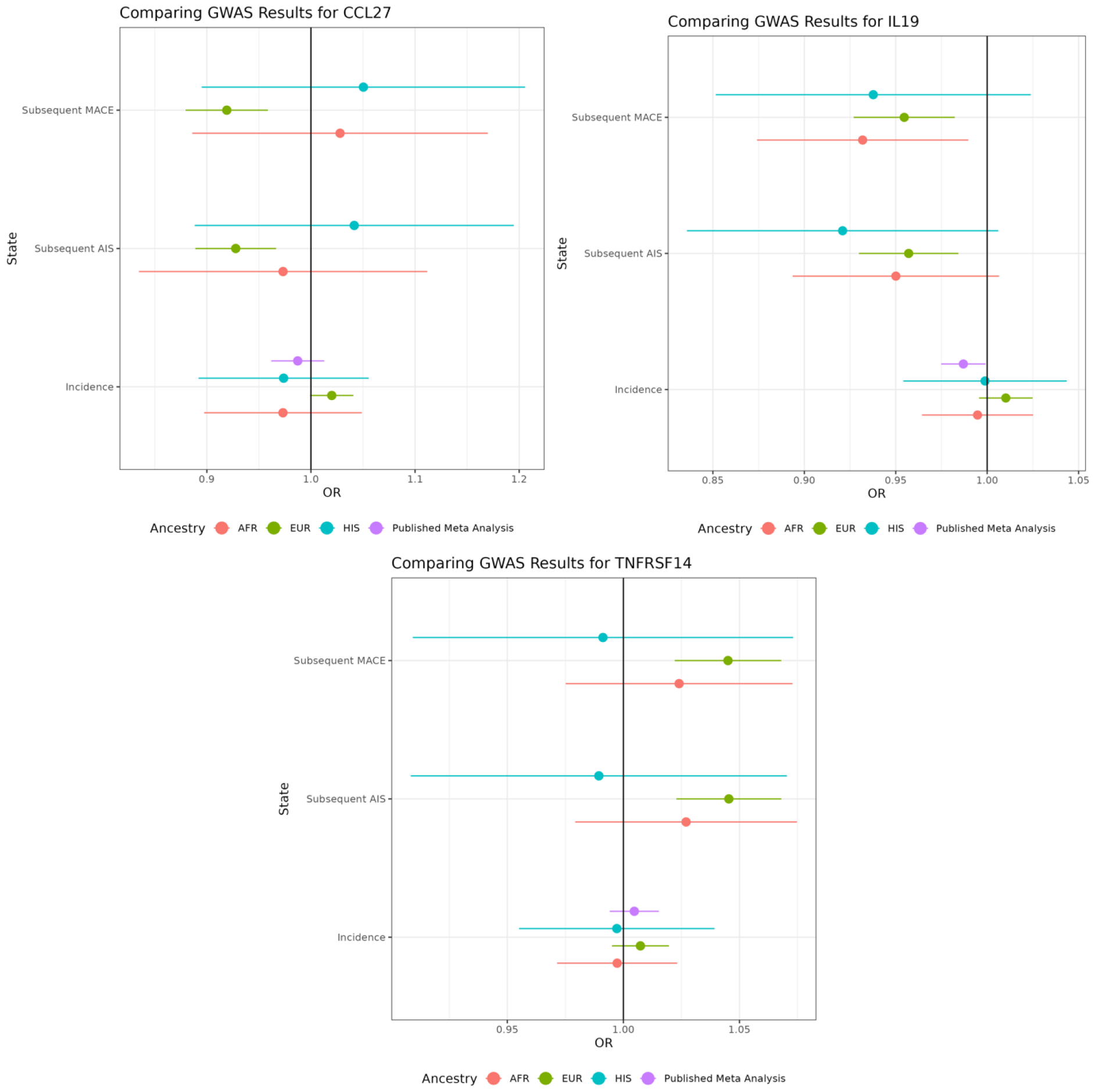
breakdown of significant MR results in subsequent stroke states by ancestry, as well as a comparison to incident stroke (Cochran Q p>=0.1). MR: Mendelian randomization; MACE: Major Acute Cardiovascular Events; AIS: Acute Ischemic Stroke; MR: Mendelian randomization; OR: Odds Ratio

#### Comparing MR Results Against Potential Druggable Targets

Of the 720 genes related to “Ischemic Stroke” in Open Targets(23), 103 had instrumentable protein products (p<1x10^-11^) in UKB-PPP(16). Five of these met an FDR-adjusted significance threshold in MR for incident stroke (ANGPT1, FGF5, FURIN, MMP12, TFPI), but none in the MR of subsequent stroke. Of the 5 targets with evidence of a causal effect on incidence, 4 were previously identified as putatively causal for incident stroke in MR studies(26,27), while FURIN is novel. ANGPT1, TFPI, and MMP12 have evidence of a causal effect on incident AIS, while FURIN and FGF5 have existing genetic associations. Using the Therapeutic Target Database(28) to find existing therapeutic drugs for these genes, we determined that TFPI and ANGPT1 have phase 3 clinical trials associated with them, MMP12 has a phase 1 trial, and FURIN has pre-clinical trial. None of the drugs which started clinical trials were designed for stroke (**Table S8**). For markers associated with subsequent MACE, currently, there are no clinical trials of drugs targeting TNFRSF14, CCL27.

## Discussion

We observed several loci associated with subsequent stroke events that have a role in inflammation. There exists a link between inflammation and stroke. While the immune response starts locally, inflammatory mediators propagate, which leads to a systemic inflammatory response, followed by immunosuppression(29). Changes in TNF and IL6 levels have been observed in patients at the onset of stroke(30). This response may be due to a state of immunodepression that occurs post-stroke, as there are increased risks of poststroke infections(29). There is increasing evidence that greater inflammation is associated with AIS progression. It is unclear whether inflammation is transitory, related to the severity of the ischemia, and the ischemia-inflammation association post stroke is not well characterized(31).

The discovery of two proteins having a predicted causal effect on subsequent MACE after stroke suggests that inflammation is a contributing factor to subsequent MACE outcomes after incident stroke AIS(24,25). TNFRSF14 (also known as HVEM) signals via TRAF2/3 pathway, role in immune cell survival. TNFRSF14 is a receptor for 4 ligands: TNFSF14 (LIGHT), LTA, BTLA and CD160. First two are TNF cytokines, 2nd two are Ig-related membrane proteins. HVEM has been shown to contribute to plaque destabilization and rupture(32). The LIGHT protein is known to have prognostic predictive value for composite cardiovascular events(33). The TNF-alpha family also has been suggested as being a risk factor to stroke(34). CD160 has been shown to be a potential indicator of the progression of atherosclerosis(35). Plasma measures of three of these four ligands were available in the UKB proteomics data, but (despite having strong instruments available F>500) neither had a causal effect on subsequent MACE (LTA p=0.634; CD160 p=0.184, TNFSF14 p=0.303). CCL27 is a cytokine involved in maintaining immune homeostasis in barrier tissues(36).

A third protein, IL19, showed slighty weaker evidence of a causal effect on subsequent MACE (OR=0.878, 95% CI = 0.81, 0.94, adj. p=0.053, coloc h4=69%) and also no effect in incident AIS (OR=0.963, 95% CI = 0.92, 1.00, adj. p=0.496). IL19 is an anti-inflammatory marker(37), and diminishes cerebral infarction and neurological deficits following cerebral ischemia in mice, potentially through the elevated expression of genes related to pro-inflammatory cytokines(38). As increased IL19 levels appear to be an anti-inflammatory marker, and increased TNFRSF14 levels show as a correlative effect as a known inflammation marker, this leads to the notion of inflammation as a contributor to subsequent MACE outcomes. As CCL27 is used in maintaining immune homeostasis in barrier tissues, is more difficult to ascertain what a negative effect size could infer without further investigation.

We observed genetic variants that appear exclusively associated with subsequent MACE and AIS after an incident AIS. This might imply novel biological insights into the disease progression of stroke. We observed that all 6 proteins that show a putatively causal effect on incident AIS do not appear to affect subsequent strokes. All individuals in this study were diagnosed, treated, and likely given blood pressure medications, statins, or both, which could mask an effect on subsequent stroke risk.

Of the 5 targets identified by MR from the drug target list in Open Targets (ANGPT1, FGF5, FURIN, MMP12, TFPI), none are associated with subsequent MACE. This suggests that these proteins may be important therapeutic targets to reduce risk for incident AIS, but not for subsequent MACE. Existing targets in Open Targets is in part populated by correlations in genetic association, thus why they initially became candidate targets. We postulate that genetic variants and genes for incident stroke are not good targets for drug discovery of subsequent stroke events.

We note that while Cochran’s Q-Statistic for heterogeneity did not show evidence for different effects between ancestries (**Table S9)**, the difference in sample sizes between ancestries remain large. Results from European ancestry were overall much stronger due to higher power. More data around individuals of non-European ancestries is necessary to investigate this further.

We had several limitations to our study. First, MACE is defined as a combination of MI, AIS/TIA and ASCVD death. For incident MACE in MVP, we have observed that MI accounts for a larger proportion of the MACE phenotype compared to AIS/TIA(39). However, our subsequent cohort has a smaller proportion of MI events than expected in the MACE phenotype **(Table S10**.) This is likely due to the selection on incident AIS. Secondly, despite analyzing relatively large datasets for disease progression, our results have limited statistical power due to sample sizes. Thirdly, as cis-only pQTL data sets are normally instrumented by a single SNP, the Wald ratio is the only available means of estimation for MR, which restricts the type of sensitivity analyses we can perform. Colocalization reduces the risk of confounding by linkage disequilibrium, as it requires the presence of a common causal variant responsible for both traits but can’t exclude potential horizontal pleiotropy, and other sources of pleiotropy could not be tested in this study. Finally, a lack of sufficient sample sizes and access to data in ancestries other than European, African, or Hispanic mean that we cannot ascertain whether these results are generalisable across all ancestries, or if there are genetic differences by ancestry. All individuals that were diagnosed with stroke will have likely been put on common preventative medication for subsequent stroke, this treatment may be altering the progression GWAS results, however no data is available to compare against individuals diagnosed with stroke but have not been treated.

## Conclusion

We observed two novel SNPs associated with subsequent stroke events that warrant further replication. We also performed MR to identify putative causal proteins for risk of subsequent MACE in stroke patients. We observed putatively causal evidence for two novel proteins (CCL27 and TNFRS14) associated with subsequent MACE risk in pQTL, suggesting that inflammation is a contributing factor to subsequent MACE outcomes after incident stroke AIS.

## Data Availability

Data is available upon request

## Non-standard Abbreviations and Acronyms

GWAS: Genome Wide Association Study
MR: Mendelian randomization
MACE: Major Adverse Cardiovascular Events
AIS: Arterial Ischemic Stroke
TIA: Transient Ischaemic Attack
SNP: Single Nucleotide Polymorphism
pQTL: Protein Quantitative Trait Loci
UKB: United Kingdom Biobank
MVP: Million Veteran Program

## Acknowledgments

We acknowledge the VA Million Veteran Program (MVP) participants. This study was supported by the National Institute for Health and Care Research Bristol Biomedical Research Centre. The views expressed are those of the author(s) and not necessarily those of the NIHR or the Department of Health and Social Care. The UK Biobank data was obtained under the UK Biobank resource application 81499. We would like to thank the participants of MVP and UKB studies.

## Disclosures

The following authors have nothing to declare: AE, NA, HJA, DCP, GH, KT, PWFW, JPC, JMG, GDS, LP, KC, GMP

TRG receives funding from Biogen and GSK for unrelated work.

AEH started working for Novo Nordisk after contributing to this manuscript.

